# Prediction of diabetic kidney disease risk using machine learning models: a population-based cohort study of Asian adults

**DOI:** 10.1101/2022.08.17.22278900

**Authors:** Charumathi Sabanayagam, Feng He, Simon Nusinovici, Jialiang Li, Cynthia Lim, Gavin Tan, Ching-Yu Cheng

## Abstract

**Background:** Machine learning (ML) techniques improve disease prediction by identifying the most relevant features in multi-dimensional data. We compared the accuracy of ML algorithms for predicting incident diabetic kidney disease (DKD).

**Methods:** We utilized longitudinal data from 1365 Chinese, Malay and Indian participants aged 40-80 years with diabetes but free of DKD who participated in the baseline and 6-year follow-up visit of the Singapore Epidemiology of Eye Diseases Study (2004-2017). Incident DKD (11.9%) was defined as an estimated glomerular filtration rate (eGFR) <60 mL/min/1.73m^2^ with at least 25% decrease in eGFR at follow-up from baseline. 339 features including participant characteristics, retinal imaging, genetic and blood metabolites were used as predictors. Performances of several ML models were compared to each other and to logic regression (LR) model based on established features of DKD (age, sex, ethnicity, duration of diabetes, systolic blood pressure, HbA1c, and body mass index) using area under the receiver operating characteristic curve (AUC).

**Results:** ML model, Elastic Net (EN) had the best AUC (95% confidence interval) of 0.851 (0.847-0.856), which was 7.0% relatively higher than by LR 0.795 (0.790-0.801). Sensitivity and specificity of EN were 88.2% and 65.9% vs. 73.0% and 72.8% by LR. The top-15 predictors included age, ethnicity, antidiabetic medication, hypertension, diabetic retinopathy, systolic blood pressure, HbA1c, eGFR and metabolites related to lipids, lipoproteins, fatty acids and ketone bodies.

**Conclusions:** Our results showed ML together with feature selection improves prediction accuracy of DKD risk in an asymptomatic stable population and identifies novel risk factors including metabolites.

**Funding Support:** This study was supported by the National Medical Research Council, NMRC/OFLCG/001/2017 and NMRC/HCSAINV/MOH-001019-00. The funders had no role in study design, data collection and analysis, decision to publish, or preparation of the manuscript.

**Conflicts of interest:** None declared.

## INTRODUCTION

Diabetes currently affects an estimated of 415 million people worldwide in 2015, and the number is expected to increase to 642 million by 2040 with the greatest increase expected in Asia, particularly India and China [1]. With the rising prevalence of diabetes and population aging, the burden of diabetic kidney disease (DKD), a leading cause of end-stage renal disease (ESRD), cardiovascular disease (CVD), and premature deaths, is also set to rise in parallel. Diabetes accounts for 30-50% of all chronic kidney disease (CKD) cases affecting 285 million people worldwide [2]. As CKD is asymptomatic till more than 50% of kidney function decline, early detection of individuals with diabetes who are at risk of developing DKD may facilitate prevention and appropriate intervention for DKD. Early identification of individuals at risk of developing CKD in type 2 diabetes is challenging. Therefore, characterization of new biomarkers is urgently needed for identifying individuals at risk of progressive decline of eGFR and timely intervention for improving outcomes in DKD.

Several risk prediction models have been developed in the past for predicting progression to end-stage renal disease, but studies predicting onset of CKD in diabetic populations are limited. These studies were focused on clinical populations utilizing data from clinical trials [3] or heterogeneous cohorts of patients with different CKD definitions [4]. Dunkler et al. showed albuminuria and estimated glomerular filtration rate (eGFR) were the key predictors and addition of demographic, clinical or laboratory variables did not improve predictive performance beyond 69% [3]. Current CKD risk prediction models developed using traditional regression models (e.g., logistic, or linear regression) perform well when there are only small or moderate numbers of variables or predictors but tend to overfit if there is a large number of variables. Machine learning methods using ‘Big data’, or multi-dimensional data may improve prediction as they have less restrictive statistical assumptions compared to traditional regression models which assume linear relationships between risk factors and the logit of the outcomes and absence of multi-collinearity among explanatory variables.

Diabetes is a metabolic disorder and metabolic changes associated with diabetes lead to glomerular hypertrophy, glomerulosclerosis, tubulointerstitial inflammation and fibrosis [5]. Several blood metabolites have been shown to be associated with DKD. Similarly, genetic abnormalities in diabetes have also been shown to increase the risk of DKD. We and several others have previously shown that retinal microvascular changes including retinopathy, vessel narrowing, or dilation and vessel tortuosity were associated with CKD [6, 7]. Integrating high-dimensional data from multiple domains including patient characteristics, clinical and ‘Omics’ data has the potential to aid in risk-stratification, prediction of future-risk besides providing insights into the pathogenesis [8]. These features may contribute to the prediction in very complicated ways, and they may not fully satisfy the requirement for a simple linear logistic model. It is thus more appropriate to consider the ML approaches for a comprehensive study.

In the current study, we aimed to evaluate the performance of a set of most common ML models for predicting 6-year risk of DKD compared to traditional logistic regression and identify important predictors of DKD in a large population-based cohort study in Singapore with multi-dimension data including imaging, metabolites and genetic biomarkers.

## METHODS

### Study population

Data for this study was derived from the Singapore Epidemiology of Eye Diseases (SEED) study, a population-based prospective study of eye diseases in 10,033 Asian adults aged 40-80 years in Singapore. The follow-up study was conducted after a median duration of 6.08 years (interquartile range: [5.56, 6.79]) with 6,762 participants. The detailed methodology of the SEED has been published elsewhere. Briefly, the name list of adults residing in the southwestern part of Singapore was provided by the Ministry of Home Affairs, and then an age-stratified random sampling procedure was conducted. A total of 3,280 Malays (2004-2007) [9], 3,400 Indians (2007-2009) and 3,353 Chinese (2009-2011) [10] participated in the baseline study with response rates of 78.7%, 75.6% and 72.8%, respectively. As all three studies followed the same methodology and were conducted in the same study clinic, we combined the three populations for the present study. For the current analysis, we included only those with diabetes defined as random glucose ≥ 11.1 mmol/L, HbA1c ≥6.5% (48 mmol/mol), self-reported anti-diabetic medication use or having been diagnosed with diabetes by a physician based on American Diabetes association recommendations. Of the 6,762 participants who attended both baseline and follow-up visit, after excluding those without diabetes (n=5,307), prevalent CKD (n=315), missing information on eGFR (n=90), final sample size for analysis was 1,365 (47.5% Indians, 27.8% Malays and 24.7% Chinese). The sample size available for each dataset after removing participants missing >10% data was between 976 and 1,364 (**Supplementary Table S1**).

This study was performed in accordance with the tenets of the Declaration of Helsinki and ethics approval was obtained from the Singapore Eye Research Institute Institutional Review Board. Written informed consent was provided by participants.

### Assessment of DKD

Incident DKD was defined as an estimated glomerular filtration rate (eGFR) <60 mL/min/1.73m^2^ with at least 25% decrease in eGFR at follow-up in participants who had eGFR>60 mL/min/1.73m^2^ at baseline. Combining change in eGFR category together with a minimal percent change ensures that small changes in eGFR, for e.g., from 61 to 59 mL/min/1.73m^2^ is not misinterpreted as incident CKD as the eGFR is <60 mL/min/1.73m^2^ [7, 11]. The reduction in eGFR at follow-up was calculated as a percentage of the baseline eGFR as (eGFR at baseline – eGFR at follow-up)/eGFR at baseline *100%. GFR was estimated from plasma creatinine using the Chronic Kidney Disease Epidemiology Collaboration (CKD-EPI) equation [12]. Blood creatinine was measured by the Jaffe method on the Beckman DXC800 analyzer calibrated to the Isotope Dilution Mass Spectrometry (IDMS) method using the National Institute of Standards and Technology (NIST) Reference material. Based on the level of eGFR, DKD severity was classified into 4 groups: eGFR ≥60 (reference representing normal/high/mild decrease in kidney function, mild-to-moderate (eGFR 45-59), moderate-to-severe (eGFR 30-44), and severe/renal failure (eGFR<30) [13].

### Variables for prediction

We evaluated 339 features such as demographic, lifestyle, socioeconomic, physical, laboratory, retinal imaging, genetic and blood metabolomics profile. The entire list of variables is presented in **Supplementary Table S2**. We organized the variables into five different domains: traditional risk factors, extended risk factors, imaging parameters, genetic parameters, and blood metabolites. For ML analysis, based on different combinations of the five domains, we tested six models (A to F): A=Traditional risk factors; B= A+ Extended risk factors; C= B+ Imaging parameters; D= B+ Genetic parameters; E= B+ Blood metabolites; F= B+ Imaging parameters+ Blood metabolites+ Genetic parameters.

### Traditional risk factors (n=7)

Age, sex, ethnicity, body mass index (BMI), systolic blood pressure (BP), duration of diabetes and HbA1c% were included as traditional risk factors.

### Extended risk factors (n=22)

Marital status, educational level, monthly income, smoking status, alcohol consumption, history of cardiovascular disease, hypertension status, diastolic BP, pulse pressure, blood glucose, total, high-density lipoprotein (HDL) and low-density lipoprotein (LDL) cholesterol levels, anti-diabetic, anti-hypertensive, and anti-cholesterol medication use were included as part of extended risk factors.

### Blood metabolites (n=223)

We quantified 228 metabolic measures from stored serum/plasma samples at baseline using a high-throughput NMR metabolomics platform (Nightingale Health, Helsinki, Finland). The metabolites included routine lipids, lipoprotein subclasses with lipid concentrations within 14 subclasses, fatty acids, amino acids, ketone bodies, and glycolysis-related metabolites. The 14 lipoprotein subclasses include six subclasses of VLDL (extremely large, very large, large, medium, small, very small), IDL, three subclasses of LDL (large, medium, small), and four subclasses of HDL (very large, large, medium, small). Lipid concentration within each lipoprotein particle included triacylglycerol, total cholesterol, non-esterified cholesterol and cholesteryl ester levels, and phospholipid concentrations [14]. Of the 228 metabolites, pyruvate, glycerol and glycine were not available in Malays. In addition, creatinine and glucose were measured as part of the blood biochemistry. After excluding these five metabolites, 223 were included under the metabolites dataset.

### Genetic parameters (n=76)

We included 76 type 2 diabetes-associated single nucleotide polymorphisms (SNPs) identified in the largest meta-analysis of type 2 diabetes genome-wide association studies by the DIAbetes Genetics Replication and Meta-analysis (DIAGRAM) consortium [15].

### Imaging parameters (n=11)

sing a semi-automated computer program (Singapore I Vessel Assessment-SIVA) we quantified retinal imaging parameters from digital retinal photographs. The parameters included retinal arteriolar and venular diameters, vessel tortuosity, branching angle, fractal dimension etc. [7]. Diabetic retinopathy (DR) was assessed by trained graders using a standard protocol [16].

### Machine learning algorithms

We tested 9 different ML algorithms including logistic regression (LR), LASSO logistic regression (LASSO), elastic net (EN), classification and regression tree (CART), random forest (RF), gradient boosting decision tree (GBDT), extreme gradient boosting (XGB), support vector machine (SVM), and naïve Bayes (NB) [17].

### Model development

We split the study samples randomly into training (80%) and test sets (20%) of equal CKD case rate by stratified sampling, with 40 random repeats of 5-fold cross-validation to evaluate the model performance. Predictive accuracy was assessed using metrics such as area under the receiver operating characteristic curve (AUC) with 95% confidence interval (CI), sensitivity and specificity calculated at the optimal cut-point (determined by Youden’s index). In preliminary analyses, testing different combinations of features (**Figure 1A to 1F**), performance of all ML models was below 0.80 in dataset D including genetic features (best AUC= 0.785 by RF) and Dataset F including all 339 features (best AUC=0.788 by XGB). Hence, we dropped these 2 datasets (D and F) from further analyses. The performance of all ML models based on AUC (IQR) in Dataset 1A-1F is shown in **Supplementary Table S3** and based on sensitivity and specificity is shown in **Supplementary Table S4**.

**Figure 1.**
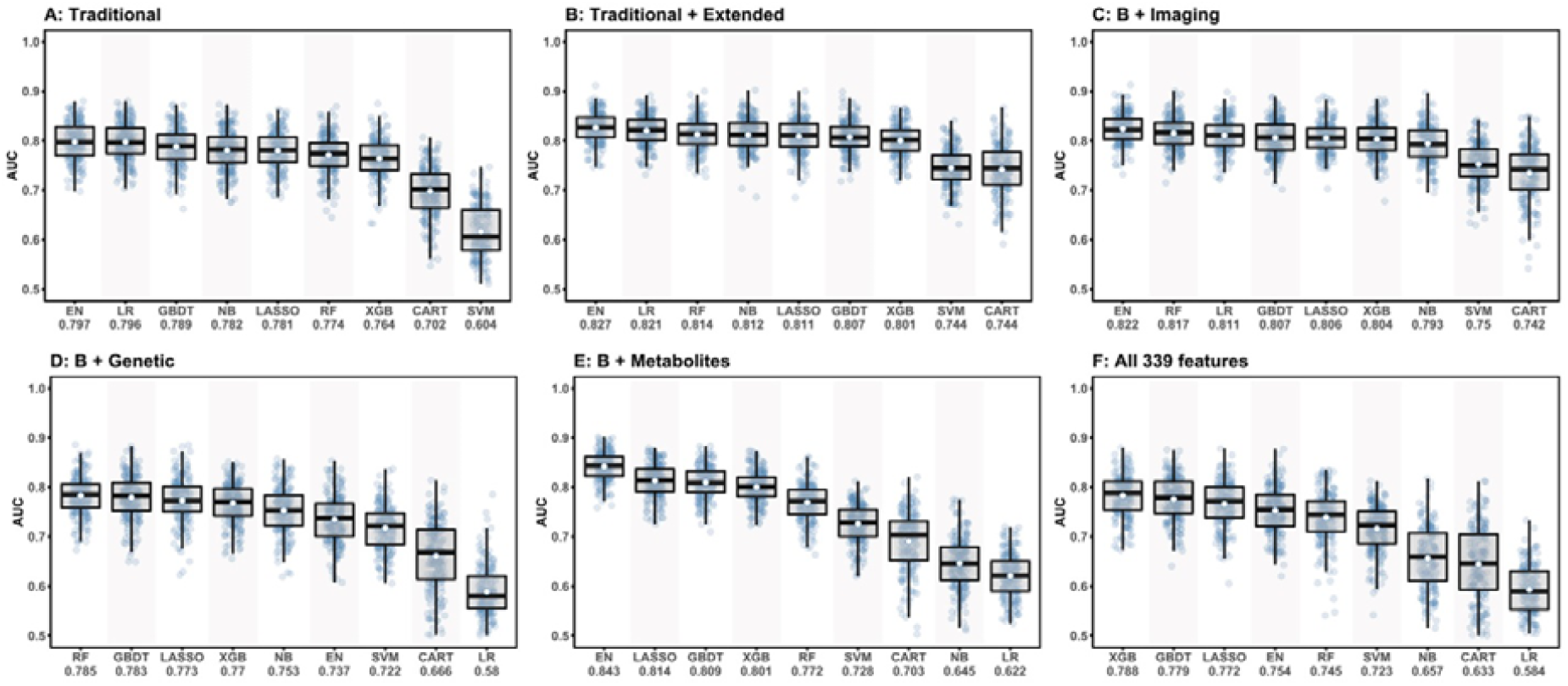
Comparison of 9 machine learning models for DKD incidence prediction.

Of the ML models, performances of CART, SVM and NB were lower compared to other models, hence these models were also dropped. Consequently, ML models EN, GBDT, LASSO, XGB and RF were considered for subsequent analyses using datasets A, B, C, and E including 252 features.

### Feature selection

All algorithms included in the current study can perform feature selection but using different selection criteria. In LR, stepwise selection according to the Akaike information criterion (AIC) is widely used but it lacks stability. LASSO is an extension of LR with L1 regularization to drop the less important variables. EN is like LASSO but with a milder regularization, resulting in a larger number of retained variables. In order to select only the most predictive features, we recursively apply EN until the retained variable subset is optimized, i.e., recursive feature selection (RFE). In RF, GBDT, and XGB, the most predictive variables were identified based on their relative importance to model performance. Feature selection was also performed according to their selection frequency during repeated cross-validation. We identified the top-15 predictors by each of the best performing ML models, then compared the performance of the ML models based on the top variables with that of logistic regression based on seven traditional risk factors (age, sex, ethnicity, BMI, HbA1c, duration of diabetes, and systolic BP) in another 40 random repeats of five-fold cross-validation.

### Statistical analyses

We compared the baseline characteristics of participants with diabetes by incident DKD status using chi-square test or Mann-Whitney U Test as appropriate for the variable. Statistical significance was defined as a p-value < 0.05. Subgroup numbers such as diabetic retinopathy status may not add up due to the presence of missing data. For modelling, we used mean values/modes for missing value imputation as appropriate for each variable because the missing proportions were all below 10%. Improvement in prediction accuracy by ML over the traditional risk factor model was calculated as (ML AUC-traditional model AUC)/traditional model AUC*100%. All analyses were conducted using R software version 4.0.2. To assess whether the features selected by ML models are meaningful, we visualized the association of top-15 variables with incident DKD in forest plots or a variable importance plot as appropriate for the algorithm.

## RESULTS

The 6-year incidence of DKD was 11.9% in the study population. Incidence of DKD was highest in Malays (18.4%), followed by Chinese (12.8%). Although Indians represent nearly half of the total diabetic population (648 of the 1365 diabetic participants, 47.5%), DKD was lowest in Indians (7.6%).

As shown in **Table 1**, compared to those without incident DKD, those with were significantly older, more likely to be Malays or Chinese, primary/below educated, had higher prevalence of hypertension, diabetic retinopathy, cardiovascular disease, anti-diabetic medication use; had longer duration of diabetes, higher levels of systolic BP and HbA1c%.

**Table 1.** Baseline characteristics of SEED Diabetic participants by incident CKD status.

| Characteristics | No CKD<br>(n = 1203) | CKD<br>(n = 162) | p-value | Overall<br>(n = 1365) |
| --- | --- | --- | --- | --- |
| Age (years) | 57.95 (8.78) | 64.63 (7.98) | <0.001 | 58.74 (8.95) |
| Gender, female | 580 (48.2) | 87 (53.7) | 0.219 | 667 (48.9) |
| Ethnicity |  |  | <0.001 |  |
| Indians (Ref) | 599 (49.8) | 49 (30.2) |  | 648 (47.5) |
| Malays | 310 (25.8) | 70 (43.2) |  | 380 (27.8) |
| Chinese | 294 (24.4) | 43 (26.5) |  | 337 (24.7) |
| Primary/below education, % | 706 (58.7) | 121 (74.7) | <0.001 | 827 (60.6) |
| Current smoker, % | 173 (14.4) | 16 (9.9) | 0.15 | 189 (13.9) |
| Alcohol consumption, % | 111 (9.2) | 11 (6.8) | 0.389 | 122 (9.0) |
| Hypertension, % | 845 (70.4) | 155 (95.7) | <0.001 | 1000 (73.4) |
| Diabetic retinopathy, % | 228 (19.2) | 56 (35.4) | <0.001 | 284 (21.1) |
| Cardiovascular disease, % | 153 (12.7) | 32 (19.8) | 0.02 | 185 (13.6) |
| Duration of diabetes (years) | 2.68 [0.00, 8.56] | 6.08 [1.44, 11.63] | <0.001 | 3.20 [0.00, 9.37] |
| Anti-diabetic medication, % | 681 (56.6) | 122 (75.3) | <0.001 | 803 (58.8) |
| Body mass index (kg/m<sup>2</sup>) | 26.96 (4.62) | 27.05 (4.36) | 0.764 | 26.97 (4.59) |
| Systolic blood pressure (mm Hg) | 139.42 (18.95) | 155.24 (20.01) | <0.001 | 141.29 (19.74) |
| Diastolic blood pressure (mm Hg) | 78.25 (9.74) | 79.14 (10.70) | 0.278 | 78.35 (9.85) |
| Random blood glucose (mmol/L) | 9.53 (4.26) | 10.44 (5.01) | 0.052 | 9.64 (4.36) |
| HbA1c, % | 7.61 (1.58) | 8.04 (1.83) | 0.003 | 7.66 (1.62) |
| Blood total Cholesterol (mmol/L) | 5.14 (1.14) | 4.98 (1.15) | 0.124 | 5.12 (1.15) |
| Blood HDL Cholesterol (mmol/L) | 1.12 (0.31) | 1.16 (0.35) | 0.178 | 1.12 (0.32) |
| eGFR (mL/min/1.73 m<sup>2</sup>) | 89.98 (14.34) | 79.40 (11.69) | <0.001 | 88.72 (14.46) |

**Abbreviations:** HDL, high-density lipoprotein cholesterol; SD, standard deviation; IQR, interquartile range.
Values for categorical variables are presented as number (percentages); values for continuous variables are given as mean (SD) or median [IQR]. p-values are given by $\chi^2$ -test or Mann-Whitney U test as appropriate for the variable.

### Performance of LR using traditional risk factors (Reference) and other domain features

The LR using the 7 traditional risk factors (age, sex, ethnicity, BMI, HbA1c, duration of diabetes, and systolic BP) had an AUC of 0.796. Performance of LR improved to 0.821 using the traditional+ extended risk factors. With additional features, performance of LR dropped significantly (AUC of 0.622 in E and 0.811 in C).

### Performance of ML models using multi-dimensional data

Using datasets, A, B, C, and E, the performances of the 5 ML models (**Figure 1A-1C and 1E**) were:

1. EN ranked first in performance in all 5 datasets with AUCs ranging from 0.797 in A to 0.843 in E
2. LASSO ranged from 0.781 in A to 0.814 in E
3. GBDT ranged from 0.789 in A to 0.809 in E
4. Performance of RF ranged from 0.772 in E to 0.817 in C
5. XGB ranged from 0.764 in A to 0.804 in C

Figure 2. shows the AUCs of the top 3 performing models. Using the top-15 predictors generated by feature selection, performance of EN improved further with an AUC (95% CI) of 0.851 (0.847-0.856), sensitivity and specificity of 88.2% and 65.9% compared to LR using seven established features with AUC of 0.795 (0.790-0.801) and sensitivity and specificity of 73.0% and 72.8%.

**Figure 2.**
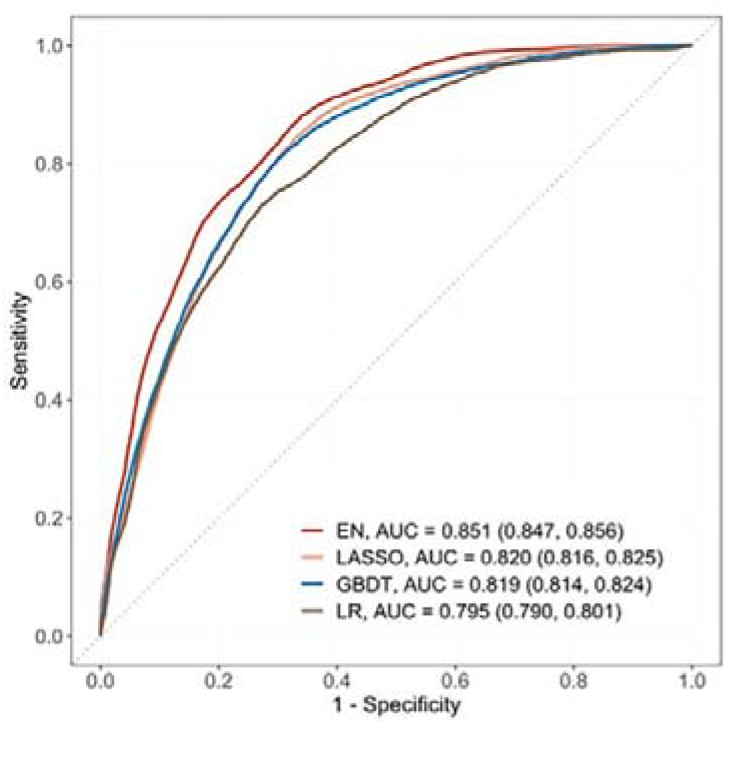
Comparison of top-3 ML models based on selected variables in dataset E (Risk factors + blood metabolites)

Corresponding estimates for LASSO were 0.820 (0.816-0.825), 84.4% and 67.0%; 0.819 (0.814-0.824), 80.6% and 70.1% for GBDT. AUC of EN, LASSO and GBDT were 7.0%, 3.1% and 3.0% relatively higher than that of LR.

### Top 15 predictors

Figure 3. shows the top 15 predictors visualised using forest plots for EN and LASSO and a variable importance plot for GBDT.

**Figure 3.**
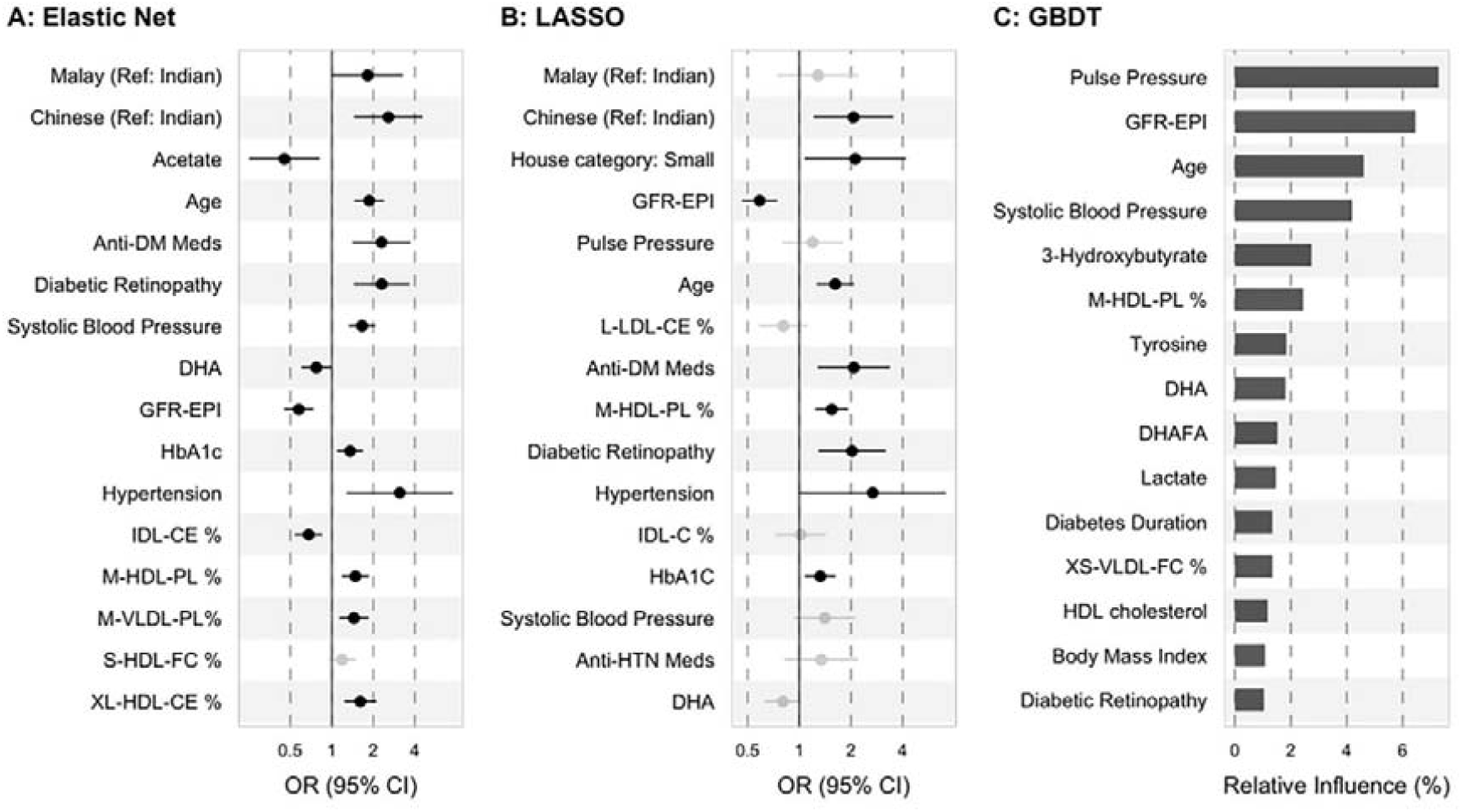
Association of top-15 ML-selected predictors with incident CKD.

Among the traditional and extended risk factors, all 3 models chose age, SBP, any diabetic retinopathy, and lower levels of eGFR as top 15 predictors. In addition, anti-diabetic medication use, HbA1c, hypertension, and ethnicity (Malay and Chinese as compared to Indians) were chosen as risk factors by EN and LASSO; anti-hypertensive medication and low housing type by LASSO; duration of diabetes, BMI and HDL cholesterol by GBDT. Among the metabolites, phospholipids to total lipids ratio in MHDL and DHA were selected by all 3 models. Free cholesterol to total lipids ratio in small HDL/XSVLDL, cholesterol esters to total lipids ratio in IDL/LLDL/XLHDL were also found of high frequency. Additionally, higher levels of acetate were shown to be protective by LR based on EN-selected variables, while tyrosine and lactate were identified as important factors by GBDT. Source data for the forest plots are shown in **Supplementary Table S5**.

## DISCUSSION

The results of the current study suggest that prediction using ML models with selected features provided improved prediction compared to LR model based on seven established features in this extensively phenotyped large-scale epidemiological study. The best performance was obtained by EN model based on dataset E including risk factors and metabolites with AUC of 0.851 which was 7.0% higher than that of LR using seven established risk factors. Sensitivity was also higher by EN (88.2% and 65.9%) compared to LR (73.0% and 72.8%). Top-15 predictors by EN using RFE identified several metabolites related to lipid concentration, lipoprotein subclasses, fatty acids, and ketone bodies as novel predictors besides confirming traditional predictors including age, ethnicity, antidiabetic medication use, presence of hypertension, diabetic retinopathy, higher levels of systolic blood pressure, HbA1c, and lower levels of eGFR. Contrary to conventional risk factors, sex, BMI, and duration of diabetes did not come in the top 15-predictors.

Our results showed that ML models combined with feature selection improved the accuracy for predicting incident DKD in high-dimensional datasets. AUC of MLs based on dataset E including metabolites (+risk factors) scored highest while the one based on Dataset D including genetic features scored lowest compared to other domain features. This finding suggests that modifiable risk factors and metabolites predict DKD risk better than genetic features. Predictive performance was best by EN, followed by LASSO and GBDT. Top-15 predictors selected by LASSO and GBDT were largely consistent to that by EN.

Few previous studies have evaluated the performance of ML models for predicting the risk of incident DKD (**Table 2**). Ravizza et al. identified seven key features (age, BMI, eGFR, concentration of creatinine, glucose, albumin and HbA1c%) by a data-driven feature selection strategy for predicting DKD using EHR data from 417,912 people with diabetes retrieved from the IBM Explorys Database and developed a random forest model in 82,912 people with diabetes retrieved from Indiana Network for Patient Care (INPC). The RF algorithm using seven prioritized key features achieved an AUC of 0.833 as compared to 0.827 by logistic regression [18].

**Table 2.** Machine learning model for predicting incident CKD in literature.

| Author, journal | Study cohort<br>Country | Study<br>population<br>Follow up | CKD<br>Definition and<br>incidence | Number of<br>predictors | ML Performance |
| --- | --- | --- | --- | --- | --- |
| Ravizza et al.[18]<br>*Nature Medicine*,<br>2019 | EHR data<br>from the IBM<br>Explorys and<br>INPC datasets, | Development<br>cohort (IBM):<br>>500,000<br>adults with | ICD 9/10 codes | 300<br>features | Based on 7 prioritized<br>features, AUC by RF =<br>**0.833** and The<br>Roche/IBM supervised |
|  | US | diabetes.<br>Validation<br>(INPC)=<br>82912 adults<br>with T2DM;<br>FU=3 years |  |  | algorithm by LR = **0.827** |
| Song X et al.[19]<br>*JMIR*, 2020 | EHR data, US<br>(2007-2017) | 14039 adults<br>with T2DM.<br>FU=1-year | eGFR<60 or<br>UACR≥30 mg/g;<br>34.1% | >3000 | GBM<br>**AUC = 0.83** |
| Huang et al.[20]<br>*Diabetes*, 2021 | KORA cohort,<br>Germany | 1838 adults<br>with<br>prediabetes<br>and T2DM<br>FU=6.5y | eGFR<60 or<br>UACR≥30 mg/g<br>at FU.<br>10.9% | 125<br>mets+14<br>clinical<br>factors | SVM, RF, Ada Boost<br>Best set: Mets- SM and<br>PC+ Age, TC, FPG,<br>eGFR, UACR, **AUC =**<br>**0.857**<br>Traditional LR using 14<br>variables, **AUC = 0.809** |
| Sabanayagam et al.<br>(2022)<br>Current study | SEED<br>population<br>data,<br>Singapore | 1365 adults<br>with diabetes.<br>FU=6 years | eGFR<60<br>+25% decline<br>in eGFR from<br>baseline | 339<br>features | EN+RFE selected 15<br>features, **AUC = 0.851 vs.**<br>**0.795** using 7 features by<br>traditional LR |

Song et al. predicted 1-year risk of DKD based on EHR data using Gradient Boosting Machine (GBM) algorithm with an AUC of 83% [19]. As the median duration of development of DKD is ∼10 years since the onset of diabetes, predicting 1 year risk may not be sufficient. Huang et al. predicted DKD risk in 1,838 adults with diabetes and prediabetes who participated in the KORA Study in Germany.Authors used ML models Support Vector Machine (SVF), RF and Ada Boost based on 14 clinical factors and 125 metabolites. The best set AUC was 0.857 which is similar to that of our model using EN (AUC=0.851).

In the current study, we observed that when the features are limited to the traditional risk factors, performance of LR was similar to that of the best ML model EN, but when number of features is huge, LR performance dropped significantly compared to the top performing ML models including EN, LASSO and GBDT. In a previous study based on the same dataset as the current study, Nusinovici et al. tested the performance of several ML models utilizing 20 risk factors alone, found that the performance of LR (AUC=0.905) was similar to that of the best ML model, GBDT (AUC=0.903) for predicting incident CKD in those with and without diabetes [21]. When a large number of features are present, ML methods may capture the complicated functional dependency of the incident CKD outcome much better than the linear approach used in LR.

The risk factors identified by the top-performing ML models (EN, LASSO, GBDT) are established risk factors such as age, ethnicity, antidiabetic medication use, presence of hypertension, diabetic retinopathy, higher levels of systolic blood pressure, HbA1c, and lower levels of eGFR. Additionally, anti-hypertensive medication use, and low housing type were identified by LASSO while BMI, duration of diabetes by GBDT. Increasing age, longer duration of diabetes, higher levels of HbA1c, systolic blood pressure/hypertension are well known risk factors of DKD. Older age, hypertension, lower eGFR, higher levels of BMI, HbA1c and antidiabetic medication use were identified to be significant risk factors for incident CKD in those with diabetes by Nelson et al. in a meta-analysis including 15 multi-national cohorts with diabetes as part of the CKD Prognosis Consortium (CKD-PC) [22]. While black ethnicity was a risk factor for CKD in the meta-analysis, in our study, we found Chinese and Malay ethnicity to be at higher risk of developing incident DKD compared to Indian ethnicity. One reason for the Indian ethnicity to be at lower risk of developing DKD could be Indian ethnicity being a high-risk group for diabetes, they may be well aware of the risk, and comply with screening, medication etc. that could reduce their risk of developing DKD. Malay ethnicity has been identified to be a high-risk group for CKD by several studies conducted in Singapore. Surprisingly, gender was not identified to be a risk factor by any of the 3 ML models. This finding is consistent to Ravizza et al. algorithm based on data-driven feature selection which did not pick up gender as one of the priority features [18].

In the current study, several new predictors from the metabolites domain were identified. We found lipid metabolites including phospholipids in HDL and VLDL subclasses, cholesterol esters, and free cholesterol in HDL subclasses were associated with increased risk of DKD while cholesterol esters in IDL to be protective against DKD. Further, higher levels of DHA, acetate and tyrosine also showed a protective association (odds ratios not shown). In the ADVANCE trial, similar to our findings, higher tyrosine levels were associated with increased risk of microvascular complications in diabetic participants. DHA, a n-3 polyunsaturated fatty acid (PUFA) has been shown to reduce renal inflammation and fibrosis and slow down the progression CKD in animal models with type 2 diabetes [23] and PUFA supplementation has been shown to reduce hyperglycemia-induced pathogenic mechanisms by its anti-inflammatory and anti-oxidant properties and to improve renal function in diabetic nephropathy patients (Liborio-Neto, meta-analysis). Consistent with our findings, higher levels short-chain fatty acid acetate, have been shown to be inversely associated with diabetic nephropathy in type 2 diabetic patients [24] and to have beneficial effects in mice models with type 2 diabetes by reducing oxidative stress and inflammation.

The strengths of our study include a multi-ethnic Asian population with long follow-up and availability of a wealth of information. Use of RFE for dimension reduction and feature selection reduced overfitting of data. ML models identify the relative importance of one domain over the other domains (like metabolite features in our study compared to genetic features) and best predictors within one domain. Our study results should be interpreted in the light of few limitations. First, our definition DKD was based on measurement of single blood creatinine both at baseline and follow-up. This would have resulted in some misclassification, but the bias would be non-differential and would be similar across both outcomes. Second albuminuria, an important predictor of DKD was not included as it was missing in a substantial number of participants. Third, external validation was not performed. Fourth, ML models are computationally intensive compared to traditional regression models.

In conclusion, in a population-based sample of multi-ethnic Asian adults, we found that EN with specific metabolites outperformed the current DKD risk prediction models using demographic and clinical variables. Our results provide evidence that combining metabolites and ML models could improve prediction accuracy for DKD and that increasing use of ML techniques may discover new risk factors for DKD. Further testing in external populations would support the validity of the model.

## Data Availability

Data Availability Statement: As the study involves human participants, the data cannot be made freely available in the manuscript, the supplemental files, or a public repository due to ethical restrictions. Nevertheless, the data are available from the Singapore Eye Research Institutional Ethics Committee for researchers who meet the criteria for access to confidential data. Interested researchers can send data access requests to the Singapore Eye Research Institute using the following email address. Processed version of the datasets are provided in Supplementary Tables S1-S5.

## Data Availability Statement

As the study involves human participants, the data cannot be made freely available in the manuscript, the supplemental files, or a public repository due to ethical restrictions. Nevertheless, the data are available from the Singapore Eye Research Institutional Ethics Committee for researchers who meet the criteria for access to confidential data. Interested researchers can send data access requests to the Singapore Eye Research Institute using the following email address.

Processed version of the datasets are provided in **Supplementary Tables S1-S5**.

